# EFFECTIVENESS OF CASIRIVIMAB-IMDEVIMAB AND SOTROVIMAB MONOCLONAL ANTIBODY TREATMENT AMONG HIGH-RISK PATIENTS WITH SARS-CoV-2 INFECTION: A REAL-WORLD EXPERIENCE

**DOI:** 10.1101/2022.12.23.22283921

**Authors:** Sangeetha Murugapandian, Ahmet B. Gungor, Mohanad Al-Obaidi, Bijin Thajudeen, Ryan C. Wong, Iyad Mansour, Tirdad T. Zangeneh, Katherine M. Johnson, Nicole E. Low-Adegbija, Ruhaniyah Alam, Elvira Gonzalez-Negrete, Burhaneddin Sandıkçı, Gaurav Gupta, Edward J. Bedrick, Turcin Saridogan, Katherine Mendoza, Bekir Tanriover

## Abstract

**BACKGROUND:** Severe acute respiratory syndrome coronavirus 2 (SARS-CoV-2) can evade neutralizing antibodies, raising concerns about the effectiveness of anti-spike monoclonal antibodies (mAb).

**METHODS:** This study reports a retrospective data analysis in Banner Health Care System. Out of 109,788 adult patients who tested positive for COVID-19, the study cohort was split into patients who received Casirivimab-Imdevimab (Cas-Imd) (N=10,836; Delta-predominant period 6/2021-11/2021) and Sotrovimab (N=998; Omicron-predominant period 12/2021-1/2022) mAb compared to propensity-matched control groups (N=10,836 and N=998), respectively. Index date was the date of mAb administration or the date of positive COVID-19 testing. The primary and secondary outcomes were the incidence of composite outcome (all-cause hospitalization and/or mortality) and ICU admission at 30-days following index date, respectively.

**RESULTS:** Compared to the propensity-matched untreated control cohort, the Cas-Imd mAb reduced the composite outcome (from 7.5% to 3.7%; difference: -3.8% [95% CI: (-4.4%, -3.2%)], p <0.01) regardless of their vaccination status, while Sotrovimab mAb did not (5.0% vs. 3.8%; difference: -1.2% [95% CI: (-3.1%, 0.7%)], p =0.22). In terms of the secondary outcome, similarly Cas-Imd mAb decreased ICU admission during the first hospitalization (from 1.5% to 0.5%; difference: -1.0% [95% CI: (-1.3%, -0.7%)], p <0.01) compared to the control group, whereas Sotrovimab mAb did not (0.9% vs. 0.6%; difference: -0.3% [95% CI: (-1.2%, 0.6%)], p =0.61). Comparing the periods, the Omicron-predominant period was associated with lower composite outcome than that during the Delta-predominant period.

**CONCLUSIONS:** Cas-Imd mAb was effective against the SARS-CoV-2 Delta variant, however sotrovimab lacked efficacy in patients with SARS-CoV-2 Omicron-predominant period.

**Key Points:** This retrospective propensity matched analysis showed that treatment with Cas-Imd mAb was effective against the SARS-CoV-2 Delta variant to reduce the all-cause hospitalization and mortality within 30 days, by contrast sotrovimab mAb utilization lacked the efficacy against SARS-CoV-2 Omicron variant.

## Introduction

The severe acute respiratory syndrome coronavirus-2 (SARS-CoV-2) has caused more than 6.3 million deaths worldwide as of June 2022, with over 1 million deaths in the United States.^1^ The genome for SARS CoV-2 encodes four structural proteins, of which the spike protein is the most important.^2^ Through its receptor-binding domain, the spike protein attaches to angiotensin converting enzyme 2 (ACE2) on the host cells resulting in virus-host cell membrane fusion and subsequent viral entry. The Anti-SARS-CoV-2 monoclonal antibody targets the receptor-binding domain and has been shown to have clinical benefits, including decreasing viral load, and limiting the severity of illness. Casirivimab-imdevimab cocktail (REGEN-COV [Cas-Imd])^3^, two neutralizing human IgG1 antibodies, and sotrovimab^4,5^ are among the few anti-spike monoclonal antibodies (mAb) authorized under the Food and Drug Administration (FDA) emergency use authorization (EUA) for post-exposure treatment of non-hospitalized mild to moderate COVID-19 patients who are at high risk of progression to severe disease. In infection due to early SARS CoV-2 variants, both Cas-Imd and sotrovimab mAb administration resulted in rapid resolution of symptoms and reduced viral load leading to reduced risk of hospitalization. Between July 2021 and November 2021, the predominant SARS-CoV-2 Delta variant (B.1.617.2) was associated with higher hospitalization rates and deaths compared to prior SARS CoV-2 variants (Alpha and Beta).^6^ By November 2021, the SARS CoV-2 Omicron variant (B.1.1.529), which causes a milder infection than the Delta variant, emerged and became the dominant variant.^7,8^ Studies have shown that both the Delta and Omicron variants can evade neutralizing antibodies, raising concerns about the effectiveness of these anti-spike monoclonal antibodies against newer variants.^9,10^

The Omicron variant, first detected in specimens from Botswana and South Africa, displayed a considerably higher number of mutations in the viral spike protein compared to previous variants.^7,11^ Since becoming the dominant variant, multiple sub-lineages have evolved that are antigenically distinct from the Omicron variant and more resistant to mAbs, including sotrovimab.^12^ The extent of evasion of humoral responses has essential consequences for the future therapeutic use of monoclonal antibodies. To date, most of the data on the effectiveness of sotrovimab on the Omicron variant is based on in-vitro studies.^7^ Further, other non-neutralizing antibody-related factors could affect the efficacy and outcomes after treatment with mAbs, which can be determined only through large-scale clinical data review. We previously reported on the effectiveness of Cas-Imd in reducing the rates of hospitalization, intensive care unit (ICU) admission, and mortality during an era of predominant Delta variant.^13^ This manuscript presents real-world data using a larger sample size and reports on the effectiveness of Cas-Imd (June 2021-November 2021) and sotrovimab (December 2021-January 2022) mAb treatments among high-risk outpatients with SARS-CoV-2 infection using propensity matched cohorts.

## Methods

### Patient Consent Statement

This study was approved by the Institutional Review Board of the University of Arizona with a waiver of patient consent given the retrospective nature of the study. The study adhered to the Strengthening the Reporting of Observational Studies in Epidemiology (STROBE) statement.

### Study Design and Data Collection

In this retrospective study, the electronic health records (EHR) of COVID-19 positive patients from the Banner Health Care System (a large nonprofit healthcare organization with 30 hospitals and several associated clinics across the western United States) were reviewed. The Banner Health Care System Monoclonal Antibody Treatment program was established in December 2020 (See Supplementary document A). A multidisciplinary team reviewed patients for eligibility for monoclonal antibody treatment, guided by the FDA EUA. During the study period, there were 22 infusion sites (for the treatment cohort) and 128 testing sites in the Banner Health Care System. Data was extracted between the study period, June 1, 2021, and January 31, 2022, and censored on February 28, 2022. All patients (N = 109,788) were in an outpatient setting when the positive COVID-19 test was reported in the system. Patients who were younger than 18 years of age (N= 16,681) and those who were in hospice care or have a do-not-resuscitate status (N= 524) were excluded from the study. The resulting cohort (N= 92,583) was split into the mAb-treated cohort (N= 11,838) and the mAb-untreated control cohort (N= 80,745). Demographic and clinical covariates of these patients were extracted from Banner EHR. The propensity matching was conducted using the MatchIt package within the R statistical software and was based on 20 comorbidities from the Charlson Comorbidity Index codes (based on International Classification of Diseases, Tenth Revision [ICD-10] codes documented in the EHR within five years preceding the patient index date) in addition to patient gender, age, race/ethnicity, vaccination status, body mass index (BMI) and time period. The resulting paired study sample had 11,834 patients in each of the treated and untreated cohorts (4 treated patients unmatched), which were further analyzed based on the mAb type utilization (Cas-Imd arm [N = 10,836] and its paired untreated arm [N = 10,836] between June 2021 and November 2021; sotrovimab arm [N = 998] and its paired untreated arm [N = 998] between December 2021 and January 2022); see Figure 1 for a flow chart illustrating the study cohort selection process.

**Figure 1.**
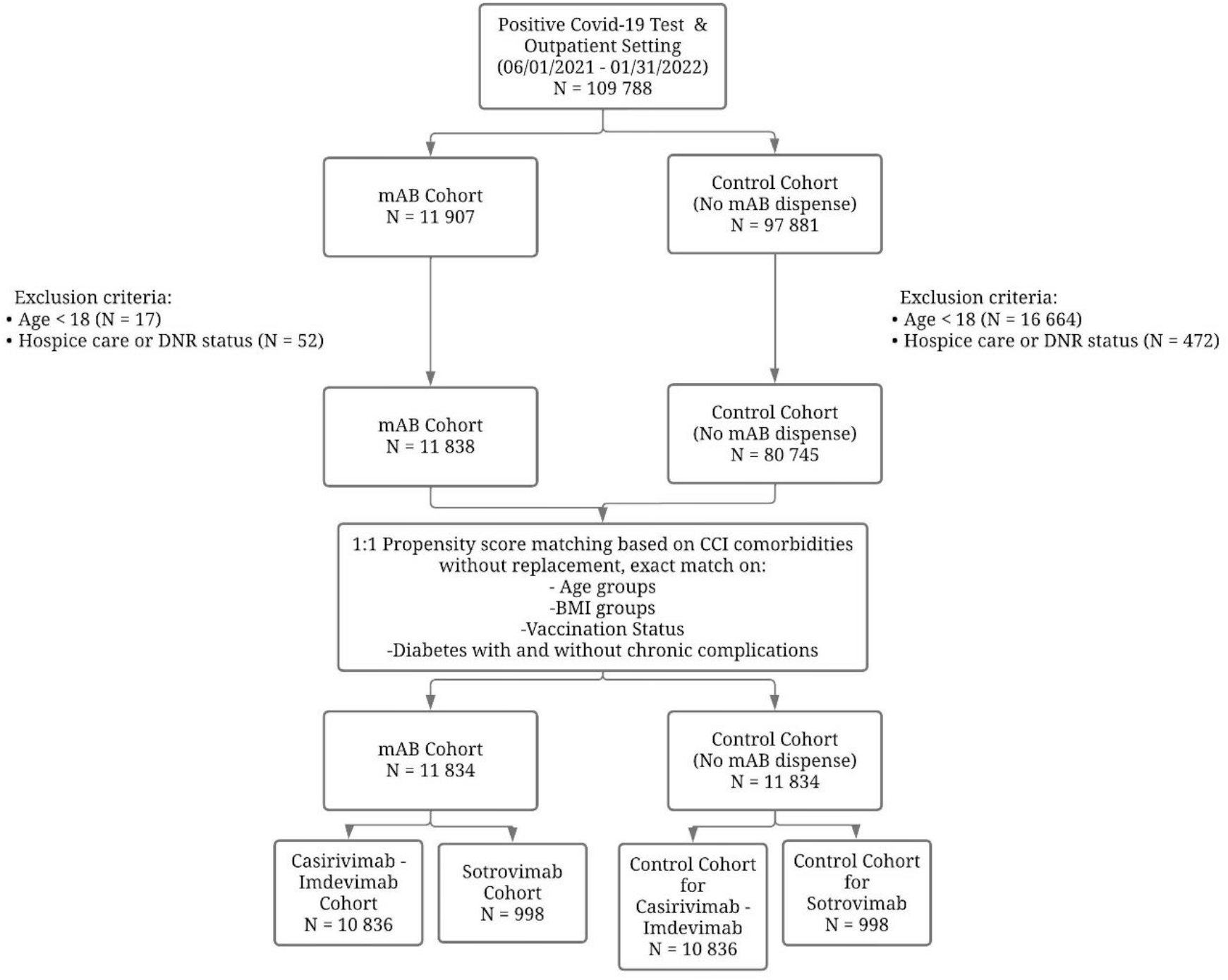
Flow chart for the study cohort selection.

### Primary and Secondary Outcomes

The index date was defined as the date mAb was received for the mAb cohort and the date of positive COVID-19 testing for the untreated control cohort. The primary and secondary outcomes were composite outcome (all-cause hospitalization and/or mortality within 30 days following the index date) and ICU admission during first hospitalization, respectively. The last follow up date was February 28, 2022. The difference in outcomes between the matched pairs were calculated using McNemar’s test. The composite outcome was further investigated among the fully vaccinated and not fully vaccinated patients in the propensity matched cohort and a subgroup analysis based on the mAb type utilization. We also reported the highest level of oxygen therapy among the post-propensity matched hospitalized patients within 30 days of the index date for the entire cohort and the subgroups separated by mAb type.

### Propensity Matching

A noted above, 26 clinical and demographic covariates were matched one-to-one using the nearest-neighbor algorithm without replacement. Pairs were exactly matched on age category, diabetes status (with and without complications), BMI categories and vaccination status (vaccinated, not fully vaccinated, unknown) to better account for the effects of these covariates on the outcomes. A patient was considered fully vaccinated only if, until their index date, 14 days have passed after their final dose of the primary series. A period variable was derived by extracting the calendar month of the index date and was also added to the propensity matching to account for possible monthly clinical differences in COVID-19 cases. The standardized mean differences (SMD) of all covariates were calculated and shown in the covariate balance plot (Figure 2). The SMD values were below the accepted 0.10 value. The improvement of SMDs from pre-match cohort to post-match cohort showed the success of propensity matching.

**Figure 2.**
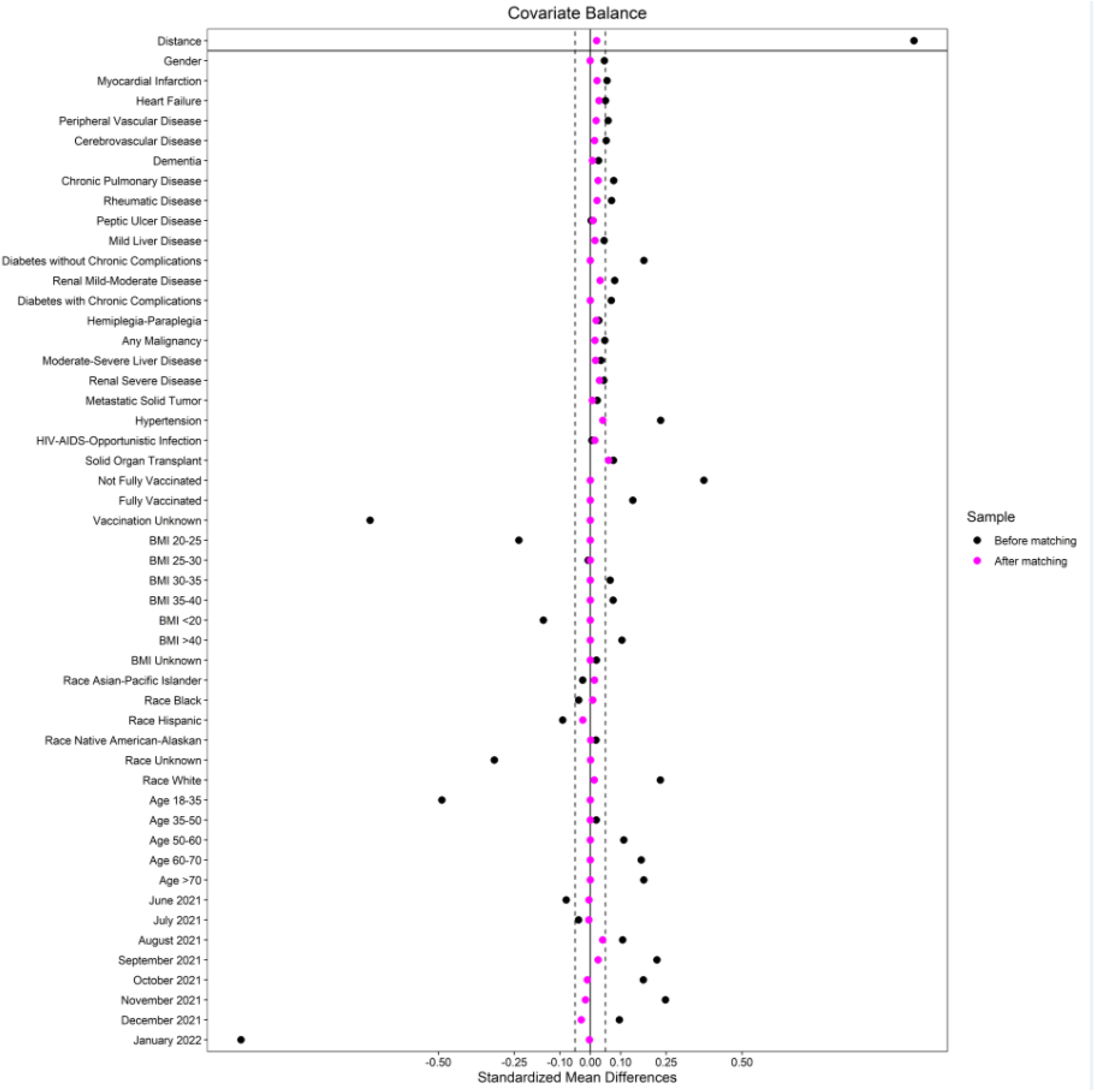
Covariate balance plot for before and after the propensity matching.

### Statistical Analysis

For each primary outcome, event counts, percentages and 95% Clopper-Pearson confidence intervals were reported. McNemar’s test using R package exact2×2 was applied to quantify the significance of differences in proportions between matched pairs, using R package exactci. The difference calculated from the test alongside the 95% confidence intervals and the p-value were reported. In addition, Kaplan-Meier survival analysis was performed to compare times to primary outcome between groups (fitted a Cox proportional hazard model accounting for the clustering defined by the pairs, results not shown) (using Stata 17, StataCorp College Station, Texas). For the intensity of oxygen therapy among the post-propensity matched hospitalized patients within 30 days of the index date, the counts, percentages, summaries for difference in proportions and p value based on a Wald test were reported for the entire study cohort and based on mAb type.

### Missing Data

Data on 3,296 (13.9% of the final study cohort) patients for vaccination status was missing.

## Results

### The Patient Characteristics

Table 1 shows the characteristics of the mAb treated and untreated cohorts before and after propensity matching. Most of the patients received their mAb within 48 hours of positive COVID-19 testing (Supplemental Figure S1-3). The sample size of the post propensity matched cohort was 23,668 and 11,834 of those patients were in the mAb treated arm. All post-propensity matching covariate SMDs were below the 0.05 threshold, indicating an acceptable matching (Figure 2). In the post-propensity matched cohort, the median age of the mAb treatment arm was 53 (interquartile range [IQR]; 40-66); 55.7% were female; 69.9% were White race; 31% were fully vaccinated and 13.9% with unknown vaccination status. Some of the well-balanced high-risk characteristics between two groups were age ≥60 (35%), BMI ≥30 (47.1%), diabetes mellitus (21.2%), hypertension (32.2%), chronic lung disease (19.8%), kidney disease (6%), combination of human immunodeficiency virus/ AIDS / opportunistic infections (5.8%), solid organ transplant (1.3%.), and any malignancy (4%). The mean ± standard deviation pulse oximetry of the participants was 97.4% ± 2.1%.

**Table 1:** Patient characteristics and covariate balance before and after propensity matching.

|  | After Propensity Matching |  |  | Before Propensity Matching |  |  |
| --- | --- | --- | --- | --- | --- | --- |
|  | mAb<br>Treatment<br>Cohort | Untreated<br>Control Cohort | SMD | mAb<br>Treatment<br>Cohort | Untreated<br>Control Cohort | SMD |
|  | N=11,834 | N=11,834 |  | N=11,838 | N=80,745 |  |
| Age | 53.0 [40.0,66.0] | 53.0 [40.0,65.0] |  | 53.0 [40.0,66.0] | 43.0 [31.0,58.0] |  |
| Age Groups |  |  |  |  |  |  |
| 18-35 | 2,004 (16.9) | 2,004 (16.9) | 0.00 | 2,004 (16.9) | 28,468 (35.3) | -0.49 |
| 35-50 | 3,373 (28.5) | 3,373 (28.5) | 0.00 | 3,373 (28.5) | 22,293 (27.6) | 0.02 |
| 50-60 | 2,315 (19.6) | 2,315 (19.6) | 0.00 | 2,315 (19.6) | 12,258 (15.2) | 0.11 |
| 60-70 | 2,184 (18.5) | 2,184 (18.5) | 0.00 | 2,188 (18.5) | 9,663 (12.0) | 0.17 |
| >70 | 1,958 (16.5) | 1,958 (16.5) | 0.00 | 1,958 (16.5) | 8,063 (10.0) | 0.17 |
| Sex |  |  |  |  |  |  |
| Male | 5,241 (44.3) | 5,244 (44.3) | 0.00 | 5,244 (44.3) | 33,906 (42.0) | 0.05 |
| Fully Vaccinated |  |  |  |  |  |  |
| Yes | 3,672 (31.0) | 3,672 (31.0) | 0.00 | 3,672 (31.0) | 19,812 (24.5) | 0.14 |
| No | 6,514 (55.0) | 6,514 (55.0) | 0.00 | 6,518 (55.1) | 29,424 (36.4) | 0.37 |
| Unknown | 1,648 (13.9) | 1,648 (13.9) | 0.00 | 1,648 (13.9) | 31,509 (39.0) | -0.73 |
| Race/Ethnicity |  |  |  |  |  |  |
| White | 8,267 (69.9) | 8,194 (69.2) | 0.01 | 8,269 (69.9) | 47,852 (59.3) | 0.23 |
| Black | 564 (4.8) | 544 (4.6) | 0.01 | 565 (4.8) | 4,507 (5.6) | -0.04 |
| Hispanic | 2,234 (18.9) | 2,348 (19.8) | -0.03 | 2,234 (18.9) | 18,103 (22.4) | -0.09 |
| Asian/Pacific Islander | 120 (1.0) | 104 (0.9) | 0.02 | 120 (1.0) | 1,022 (1.3) | -0.03 |
| Native American/Alaskan | 175 (1.5) | 173 (1.5) | 0.00 | 175 (1.5) | 1,012 (1.3) | 0.02 |
| Unknown | 474 (4.0) | 471 (4.0) | 0.00 | 475 (4.0) | 8,249 (10.2) | -0.32 |
| BMI Group |  |  |  |  |  |  |
| ≤20 | 188 (1.6) | 188 (1.6) | 0.00 | 188 (1.6) | 2,843 (3.5) | -0.15 |
| 20-25 | 1,226 (10.4) | 1,226 (10.4) | 0.00 | 1,227 (10.4) | 14,154 (17.5) | -0.24 |
| 25-30 | 2,904 (24.5) | 2,904 (24.5) | 0.00 | 2,905 (24.5) | 20,057 (24.8) | -0.01 |
| 30-35 | 2,606 (22.0) | 2,606 (22.0) | 0.00 | 2,606 (22.0) | 15,585 (19.3) | 0.07 |
| 35-40 | 1,528 (12.9) | 1,528 (12.9) | 0.00 | 1,528 (12.9) | 8,388 (10.4) | 0.08 |
| >40 | 1,449 (12.2) | 1,449 (12.2) | 0.00 | 1,449 (12.2) | 7,127 (8.8) | 0.10 |
| Unknown | 1,933 (16.3) | 1,933 (16.3) | 0.00 | 1,935 (16.3) | 12,591 (15.6) | 0.02 |
| Time period |  |  |  |  |  |  |
| 6/01-30/2021 | 133 (1.1) | 138 (1.2) | 0.00 | 134 (1.1) | 1,592 (2.0) | -0.08 |
| 7/01-31/2021 | 383 (3.2) | 392 (3.3) | 0.00 | 383 (3.2) | 3,165 (3.9) | -0.04 |
| 8/01-31/2021 | 1,538 (13.0) | 1,375 (11.6) | 0.04 | 1,538 (13.0) | 7,590 (9.4) | 0.11 |
| 9/01-30/2021 | 2,058 (17.4) | 1,942 (16.4) | 0.03 | 2,059 (17.4) | 7,320 (9.1) | 0.22 |
| 10/01-31/2021 | 1,856 (15.7) | 1,897 (16.0) | -0.01 | 1,856 (15.7) | 7,521 (9.3) | 0.18 |
| 11/01-30/2021 | 2,741 (23.2) | 2,820 (23.8) | -0.02 | 2,741 (23.2) | 10,263 (12.7) | 0.25 |
| 12/01-31/2021 | 2,197 (18.6) | 2,334 (19.7) | -0.03 | 2,199 (18.6) | 11,990 (14.8) | 0.10 |
| 01/01-30/2022 | 928 (7.8) | 936 (7.9) | 0.00 | 928 (7.8) | 31,304 (38.8) | -1.15 |

Data are presented as mean [SD] for continuous measures, and n (%) for categorical measures.

Table 1: Patient characteristics and covariate balance before and after propensity matching (continued).
|  | After Propensity Matching |  |  | Before Propensity Matching |  |  |
| --- | --- | --- | --- | --- | --- | --- |
|  | mAb<br>Treatment<br>Cohort | Untreated<br>Control<br>Cohort | SMD | mAb<br>Treatment<br>Cohort | Untreated<br>Control<br>Cohort | SMD |
|  | N=11,834 | N=11,834 |  | N=11,838 | N=80,745 |  |
| Myocardial Infarction | 359 (3.0) | 313 (2.6) | 0.02 | 360 (3.0) | 1688 (2.1) | 0.06 |
| Heart Failure | 432 (3.7) | 368 (3.1) | 0.03 | 432 (3.6) | 2185 (2.7) | 0.05 |
| Cerebrovascular Disease | 367 (3.1) | 338 (2.9) | 0.01 | 369 (3.1) | 1780 (2.2) | 0.05 |
| Hemiplegia or Paraplegia | 83 (0.7) | 64 (0.5) | 0.02 | 83 (0.7) | 376 (0.5) | 0.03 |
| Peripheral Vascular Disease | 378 (3.2) | 338 (2.9) | 0.02 | 379 (3.2) | 1744 (2.2) | 0.06 |
| Chronic Pulmonary Disease | 2341 (19.8) | 2220 (18.8) | 0.02 | 2342 (19.8) | 13490 (16.7) | 0.08 |
| Dementia | 135 (1.1) | 126 (1.1) | 0.00 | 135 (1.1) | 689 (0.9) | 0.03 |
| Hypertension | 3818 (32.3) | 3590 (30.3) | 0.04 | 3822 (32.3) | 17316 (21.4) | 0.23 |
| Diabetes without Chronic Complications | 1962 (16.6) | 1962 (16.6) | 0.00 | 1964 (16.6) | 8091 (10.0) | 0.18 |
| Diabetes with Chronic Complications | 540 (4.6) | 540 (4.6) | 0.00 | 544 (4.6) | 2539 (3.1) | 0.07 |
| Renal Mild-Moderate-Advanced Disease (CKD stage 1-4) | 565 (4.8) | 483 (4.1) | 0.03 | 566 (4.8) | 2476 (3.1) | 0.08 |
| Renal Severe Disease (CKD stage 5 and ESRD) | 137 (1.2) | 99 (0.8) | 0.03 | 138 (1.2) | 556 (0.7) | 0.04 |
| Mild Liver Disease | 630 (5.3) | 588 (5.0) | 0.02 | 630 (5.3) | 3469 (4.3) | 0.05 |
| Moderate to Severe Liver Disease | 135 (1.1) | 113 (1.0) | 0.02 | 135 (1.1) | 620 (0.8) | 0.04 |
| Peptic Ulcer Disease | 95 (0.8) | 85 (0.7) | 0.01 | 95 (0.8) | 628 (0.8) | 0.00 |
| Rheumatic Disease | 363 (3.1) | 317 (2.7) | 0.02 | 363 (3.1) | 1500 (1.9) | 0.07 |
| Malignancy, skin cancers and lymphoproliferative disorders | 372 (3.1) | 340 (2.9) | 0.02 | 372 (3.1) | 1865 (2.3) | 0.05 |
| Metastatic Solid Tumor | 95 (0.8) | 88 (0.7) | 0.01 | 95 (0.8) | 482 (0.6) | 0.02 |
| HIV/AIDS/Opportunistic Infections | 683 (5.8) | 641 (5.4) | 0.02 | 683 (5.8) | 4562 (5.6) | 0.01 |
| Solid Organ Transplant | 153 (1.3) | 72 (0.6) | 0.06 | 153 (1.3) | 348 (0.4) | 0.08 |

Data are presented as mean [SD] for continuous measures, and n (%) for categorical measures.
Abbreviations: mAb= monoclonal antibody; SMD= standardized mean difference; IQR= interquartile range; BMI= body mass index; CKD= chronic kidney disease; ESRD= end-stage renal disease; HIV= Human Immunodeficiency Virus; AIDS= acquired immunodeficiency syndrome.

### The Primary and Secondary Outcomes

Between the dates of 6/1/2021 and 1/31/2022, COVID-19 positive individuals who received mAb experienced a lower risk of the primary composite outcomes compared to post-propensity matched untreated controls (Table 2 and Supplemental Figure S4-6). Overall, the effect of COVID-19 mAb administration showed a significant reduction in the primary composite outcome (3.7%) compared to post-propensity score-matched control (7.3%) (difference [95% CI] -3.6% (-4.1%, -3.0%), p <0.01). mAb reduced 30-day all-cause hospitalization by 3.5% (95% CI (-4.0%, -2.9%), p <0.01) and 30-day mortality by 0.8% (95% CI (-1.0%, -0.6%), p <0.01) (Table 2). In terms of the secondary outcome, mAb administration showed significant reduction in ICU admission during the first hospitalization. (Difference [95% CI] -0.9% [-1.2%, -0.7%], p <0.01) (Table 2).

**Table 2:** The primary and secondary outcomes in the post-propensity score-matched cohorts.

| Primary outcomes in post-propensity score-matched cohorts |  |  |  |  |  |  |
| --- | --- | --- | --- | --- | --- | --- |
|  | mAb Treatment Cohort |  | Untreated Control Cohort |  | Difference in % with 95% CI** | P-value |
|  | N (%) | 95% CI* | N (%) | 95% CI* |  |  |
| Composite outcome within 30 days |  |  |  |  |  |  |
| Whole cohort | 441 (3.7) | 3.4, 4.1 | 862 (7.3) | 6.8, 7.8 | -3.6 (-4.1, -3.0) | <0.01 |
| Cas-Imd | 403 (3.7) | 3.4, 4.1 | 812 (7.5) | 7.0, 8.0 | -3.8 (-4.4, -3.2) | <0.01 |
| Sotrovimab | 38 (3.8) | 2.7, 5.2 | 50 (5.0) | 3.7, 6.6 | -1.2 (-3.1, 0.7) | 0.22 |
| All-cause hospitalization within 30 days |  |  |  |  |  |  |
| Whole cohort | 439 (3.7) | 3.4, 4.1 | 849 (7.2) | 6.7, 7.7 | -3.5 (-4.0, -2.9) | <0.01 |
| Cas-Imd | 401 (3.7) | 3.4, 4.1 | 800 (7.4) | 6.9, 7.9 | -3.7 (-4.3, -3.1) | <0.01 |
| Sotrovimab | 38 (3.8) | 2.7, 5.2 | 49 (4.9) | 3.7, 6.4 | -1.1 (-3.0, 0.7) | 0.26 |
| Mortality within 30 days |  |  |  |  |  |  |
| Whole cohort | 18 (0.2) | 0.1, 0.2 | 111 (0.9) | 0.8, 1.1 | -0.8 (-1.0, -0.6) | <0.01 |
| Cas-Imd | 18 (0.2) | 0.1, 0.3 | 106 (1.0) | 0.8, 1.2 | -0.8 (-1.0, -0.6) | <0.01 |
| Sotrovimab | 0 (0.0) | 0.0, 0.4 | 5 (0.5) | 0.2, 1.2 | -0.5 (-1.2, 0.0) | 0.06 |
| Secondary outcome in post-propensity score-matched cohort |  |  |  |  |  |  |
|  | mAb Treatment Cohort |  | Untreated Control Cohort |  | Difference in % with 95% CI** | P-value |
|  | N (%) | 95% CI* | N (%) | 95% CI* |  |  |
| ICU admission during first hospitalization |  |  |  |  |  |  |
| Whole cohort | 57 (0.5) | 0.4, 0.6 | 167 (1.4) | 1.2, 1.6 | -0.9 (-1.2, -0.7) | <0.01 |
| Cas-Imd | 51 (0.5) | 0.4, 0.6 | 158 (1.5) | 1.2, 1.7 | -1.0 (-1.3, -0.7) | <0.01 |
| Sotrovimab | 6 (0.6) | 0.2, 1.3 | 9 (0.9) | 0.4, 1.7 | -0.3 (-1.2, 0.6) | 0.61 |

Abbreviations: mAb= monoclonal antibody; Cas-Imd= Casirivimab-Imdevimab; CI= Confidence Interval; ICU= intensive care unit.
\* The Clopper-Pearson method was used to calculate 95% confidence intervals for the outcome percentages using the R package (Exactci).
\*\* CI for difference in paired proportions between the treatment and control cohorts.

### Subgroup Analysis Stratified by Monoclonal Antibody Type and COVID-19 Vaccination Status

Table 3 shows the primary outcomes for COVID-19 positive individuals stratified by mAb type and COVID-19 vaccination status. Patients who were not fully vaccinated and received mAb showed a significant reduction in their risk of primary outcome compared to those who did not receive mAb treatment (5.6% vs. 11.0%, difference: -5.4%, 95% CI: (-6.3%, -4.4%), p <0.01). Vaccinated individuals who received mAb also showed a significant reduction compared to their matched controls (1.7% vs. 3.6%, difference: -1.8%, 95% CI: (-2.6%, -1.1%), p <0.01). Irrespective of patients’ vaccination status, Cas-Imd resulted in a significantly lower risk of all-cause mortality and/or hospitalization within 30 days of index date compared to matched untreated controls, while sotrovimab recipients did not experience a significant difference in their rates of the primary composite outcome compared to their matched untreated controls (Table 3).

**Table 3:** The primary composite outcome stratified by patient vaccination status* among the propensity matched study cohort.

| **Primary outcome in the post-propensity-matched study cohort with known vaccination status** |  |  |  |  |  |  |
| --- | --- | --- | --- | --- | --- | --- |
|  | mAb Treatment Cohort |  | Untreated Control Cohort |  | Difference in % with 95% CI*** | *P-value* |
|  | N (%) | 95% CI* | N (%) | 95% CI** |  |  |
| Fully vaccinated<br>N=3,672 | 64 (1.7) | 1.3, 2.2 | 131 (3.6) | 3.0, 4.2 | -1.8 (-2.6, -1.1) | <0.01 |
| Not fully vaccinated<br>N=6,514 | 366 (5.6) | 5.1, 6.2 | 717 (11.0) | 10.3, 11.8 | -5.4 (-6.3, -4.4) | <0.01 |

| **Primary outcome in the post-propensity-matched casirivimab-imdevimab and untreated control cohort with known vaccination status** |  |  |  |  |  |  |
| --- | --- | --- | --- | --- | --- | --- |
|  | Casirivimab-Imdevimab Treatment Cohort |  | Untreated Control Cohort |  | Difference in % with 95% CI*** | *P-value* |
|  | N (%) | 95% CI* | N (%) | 95% CI** |  |  |
| Fully vaccinated<br>N=3,213 | 51 (1.6) | 1.2, 2.1 | 116 (3.6) | 3.0, 4.3 | -2.0 (-2.8, -1.2) | <0.01 |
| Not fully vaccinated<br>N=6,062 | 342 (5.6) | 5.1, 6.3 | 684 (11.3) | 10.5, 12.1 | -5.6 (-6.6, -4.6) | <0.01 |

| **Primary outcome in the post-propensity-matched sotrovimab and untreated control cohort with known vaccination status** |  |  |  |  |  |  |
| --- | --- | --- | --- | --- | --- | --- |
|  | Sotrovimab Treatment Cohort |  | Untreated Control Cohort |  | Difference in % with 95% CI*** | *P-value* |
|  | N (%) | 95% CI* | N (%) | 95% CI** |  |  |
| Fully vaccinated<br>N=459 | 13 (2.8) | 1.5, 4.8 | 15 (3.3) | 1.8, 5.3 | -0.4 (-3.0, 2.1) | 0.85 |
| Not fully vaccinated<br>N=452 | 24 (5.3) | 3.4, 7.8 | 33 (7.3) | 5.1, 10.1 | -2.0 (-5.3, 1.3) | 0.25 |

Abbreviations: mAb= monoclonal antibody; CI= Confidence Interval.
\*This analysis included the patients with known vaccination status only.
\*\*The Clopper-Pearson method was used to calculate 95% confidence intervals for the outcome percentages using the R package (Exactci).
\*\*\* CI for difference in paired proportions between the treatment and control cohorts.

Among the individuals who required hospital admission within 30 days of a positive COVID-19 test, the mAb administration resulted in less frequent supplemental O_2_ use (p <0.01) and decreased risk of ICU admission (p < 0.01) (Supplemental Table S1). When examining specific monoclonal antibody, Cas-Imd reduced risk of supplemental O_2_ requirement (p <0.001) and admission to ICU (p <0.01), but sotrovimab did not (p = 0.67 and 0.98, respectively). Notably, vaccination status in individuals requiring hospitalization within 30 days were comparable between the groups.

## Discussion

This study presents the real-world effect of two COVID-19 mAbs (Cas-Imd and sotrovimab) against different SARS-CoV-2 variants, using a propensity-matched cohort, over a period of eight months of the COVID-19 pandemic. Results indicated that the combined mAbs use was associated with a reduction in hospitalizations and mortality within 30 days from infection. This effect, however, is mAb specific: composite outcome of death, hospitalization or ICU admission within 30 days is found significant for Cas-Imd but not for sotrovimab.

In two previous clinical trials, Cas-Imd and sotrovimab were shown to reduce hospitalization or death among high-risk COVID-19 patients.^3,4^ However, both trials were conducted during different periods of circulating SARS-CoV-2 variants which were more susceptible to Cas-Imd and sotrovimab. Since then, new variants have emerged that have reduced the efficacy of seven mAbs (including etesevimab, bamlanivimab, imdevimab, casirivimab, tixagevimab, cilgavimab, and sotrovimab).^14,15^ This has led the FDA to discontinue EUA of some of the mAbs due to their reduced efficacy.^16^

In a previous publication, this healthcare system’s real-world experience with Cas-Imd in high-risk COVID-19 patients indicated that Cas-Imd use led to a reduced number of hospitalizations, ICU admissions, and overall mortality.^13^ In that study, Cas-Imd was used during the SARS-CoV-2 Delta variant period where Cas-Imd retained efficacy. However, Cas-Imd EUA was subsequently withdrawn for its lack of effectiveness against Omicron variants.^17^ In this paper, the analysis is expanded by including a larger population from the hospital network (Banner Healthcare System) to evaluate the impact of different COVID-19 mAbs in non-hospitalized high-risk patients with COVID-19. Therefore, both efficacy of sotrovimab and Cas-Imd, whose use was continued due to its retained efficacy during the Delta and Omicron (BA.1) variant periods, were evaluated.^18^ In contrast to previous findings, the measurable combined mAbs effect, resulted only in reduced hospitalizations within 30 days with a loss of statistically significant reduction in all-cause 30 days mortality. However, when each mAb was considered separately, it was found that Cas-Imd retained its efficacy in reducing both all-cause 30 days mortality and hospitalizations. This effect was not observed in the sotrovimab group. Hence, in contrast to the sotrovimab clinical trial, no statistically significant effect of sotrovimab in reducing all-composite outcomes, death, hospitalizations, or ICU admissions was found.^4,5^ The lack of statistical significance was also observed in the level of oxygen requirements for hospitalized patients treated with sotrovimab when compared with the control cohort in 30 days. This may be related in part to the increased immunization rates during the later period of sotrovimab use, as compared to the earlier Cas-Imd cohort. While this may explain the lack of sotrovimab effect, it may not explain the lack of sotrovimab efficacy, as the efficacy of current COVID-19 vaccines against the Omicron variants is lower than that against the Delta variant, and tends to wane faster over the months following immunization.^19^.

Unfortunately, this cohort did not allow to measure the neutralizing antibodies against different SARS-CoV-2 variants to evaluate the impact of population immunity on the efficacy of mAbs. Another explanation is that Omicron variants have lower virulence than the Delta variant, which could have reduced the effect of sotrovimab in this cohort. This is due to the numbers observed in both control and sotrovimab arms, each of which had lower all-composite outcomes compared to the Cas-Imd cohort.

Such observations were reported in previous epidemiological studies, which suggest that while the Omicron variant significantly increased transmissibility, it was less virulent, resulting in lower rates of hospital admissions and deaths.^20,21^ Another possible explanation is that increased immunity from natural infections with previous variants could have reduced hospitalizations, but studies evaluating the efficacy of neutralizing antibodies produced from previous infections failed to show efficacy against the Omicron variant.^22^

This study is not without limitations. There was a lack of baseline neutralizing antibody measurements that could help identify patients with immunity against different SARS-CoV2, blunting the effect of mAb. While this study included a large cohort, the retrospective design suffers from the limitations of unmeasured confounding factors. Finally, this study cohort’s immunization rate was lower than what was previously reported by the Arizona Department of Health Services (AZDHS).^23^

Some study strengths include the use of one of the largest and most diverse patient population cohorts ever reported; conducting a propensity-matched cohort to limit confounding factors; and the fact that data collection reflect the real-world utility of mAb against COVID-19 during a period of different immunization rates and the emergence of new variants.

To conclude, this study expands on a previous report on the real-world effect of Cas-Imd and sotrovimab against different SARS-CoV-2 variants over eight months of the COVID-19 pandemic in a large diverse patient population cohort. Combined mAbs use was associated with a reduction of hospitalizations within 30 days from infection; however, this effect was mAb specific and all-composite outcome of death, hospitalization, and ICU admission within 30 days were significant in the Cas-Imd group, but not in the sotrovimab group. Further studies are required to evaluate the causes that led to the lack of efficacy in the sotrovimab group as compared to the Cas-Imd group.

## Supporting information

STROBE statement

Disclosure form

## Data Availability

All data produced in the present study are available upon reasonable request to the authors.

## Acknowledgment

The authors thank the Banner University Medical Group – Tucson CEO, Chad Whelan, MD, Physician Executive Joshua Lee, MD, and CMO Gordon Carr, MD, for their support.

## Data Sharing Statement

The data that support the findings of this study are available on request from the corresponding author. The data are not publicly available due to privacy or ethical restrictions.

## Supplemental Document

Supplemental Document A: Website for the Banner Health COVID-19 Treatment.

## Supplemental Table

Supplemental Table S1. The highest level of oxygen support therapy among the post-propensity matched hospitalized patients within 30 days of the index date.

## Supplemental Figures

Supplemental Figure S1. Duration between COVID-19 test positivity and monoclonal antibody infusion among casirivimab-imdevimab and sotrovimab cohorts (days).

Supplemental Figure S2. Duration between COVID-19 test positivity and monoclonal antibody infusion among casirivimab-imdevimab monoclonal antibody cohort (days).

Supplemental Figure 3. Duration between COVID-19 test positivity and monoclonal antibody infusion among sotrovimab monoclonal antibody cohort (days).

Supplemental Figure S4. Kaplan-Meier survival curves for the effect of casirivimab-imdevimab and sotrovimab monoclonal antibodies administration on the composite outcome compared to the post-propensity matched untreated control cohort.

Supplemental Figure S5. Kaplan-Meier survival curves for the effect of casirivimab-imdevimab monoclonal antibody administration on the composite outcome compared to the post-propensity matched untreated control cohort.

Supplemental Figure S6. Kaplan-Meier survival curves for the effect of sotrovimab monoclonal antibody administration on the composite outcome compared to the post-propensity matched untreated control cohort.

