## Supplementary material for "EFFECTIVENESS OF CASIRIVIMAB-IMDEVIMAB AND SOTROVIMAB MONOCLONAL ANTIBODY TREATMENT AMONG HIGH-RISK PATIENTS WITH SARS-CoV-2 INFECTION: A REAL-WORLD EXPERIENCE": STROBE statement

### Modified STROBE Statement—checklist of items that should be included in reports of observational studies (Cohort Sstudies)

| | Item No | Recommendation | Authors' Statement |
|---|---|---|---|
| **Title and abstract** | 1 | (*a*) Indicate the study's design with a commonly used term in the title or the abstract | (Abstract) "a retrospective cohort study regarding comparative effectiveness of casirivimab-Imdevimab (Cas-Imd) and Sotrovimab monoclonal antibody (MAb) use" |
| | | (*b*) Provide in the abstract an informative and balanced summary of what was done and what was found | (Abstract) "counts, percentages, and confidence intervals are provided; the Sotrovimab MAb use lacked efficacy in patients with SARS-CoV-2 Omicron subvariants compared to control; while Cas-Imd MAb use proved useful in decreasing outcomes with Delta subvariants" |
| **Introduction** | | | |
| Background/rationale | 2 | Explain the scientific background and rationale for the investigation being reported | (Intro) describes lack of Phase III trials and real world experiences regarding effectiveness of MAbs in era of SARS-CoV-2 Omicron and Delta subvariants. |
| Objectives | 3 | State specific objectives, including any prespecified hypotheses | (Intro) "Comparison of all-cause hospitalization and/or death over 30-day in high-risk outpatients, who received MAb compared to the propensity score (PS) matched untreated control group for COVID-19 dominated by SARS-CoV-2 Omicron and delta subvariants" |
| **Methods** | | | |
| Study design | 4 | Present key elements of study design early in the paper | 1:1 propensity matched without replacement across 26 covariates using an optimal matching algorithm |
| Setting | 5 | Describe the setting, locations, and relevant dates, including periods of recruitment, exposure, follow-up, and data collection | Method/Overview "This observational retrospective cohort study of positive COVID-19 patients was conducted between June 1, 2021, and January 31, 2022. Patients' follow-up date was censored on February 28, 2022. All data pertaining to MAb treated patients and untreated patients were captured from electronic health records (Cerner EHR) in the Banner Health Care System, which houses thirty hospitals and several clinics across the Southwestern United States, mainly in Arizona ." |

| | | | |
|---|---|---|---|
| Participants | 6 | (*a*) *Cohort study*—Give the eligibility criteria, and the sources and methods of selection of participants. Describe methods of follow-up<br><br>*Case-control study*—Give the eligibility criteria, and the sources and methods of case ascertainment and control selection. Give the rationale for the choice of cases and controls<br><br>*Cross-sectional study*—Give the eligibility criteria, and the sources and methods of selection of participants | Cohort study, as described in Methods, Flow Chart (Figure 1), and follow-up summary (Table 1). |
| Variables | 7 | Clearly define all outcomes, exposures, predictors, potential confounders, and effect modifiers. Give diagnostic criteria, if applicable | All-cause hospitalization and death within 30 days of index date, as detailed in methods. The exposure variable (MAb use) clearly called out. The Cas-Imd and Sotrovimab MAb and control group were propensity matched based on 26 variables in the EHR, explained in Methods, the covariant balance pre and post-PS match was assessed (Table 1). The study conserved sample size/power for main effect estimation; effect modification was not performed. |
| Data sources/ measurement | 8* | For each variable of interest, give sources of data and details of methods of assessment (measurement). | Clinical covariates were derived from the Charlson Comorbidity Index codes (based on International Classification of Diseases, Tenth Revision [ICD-10] codes) documented in the Cerner-EHR within five years preceding the index date. |
| Bias | 9 | Describe any efforts to address potential sources of bias | To reduce selection bias associated with the decision to administer MAb, we utilized a 1:1 propensity matched without replacement across 26 covariates using an optimal matching algorithm that minimizes the sum of absolute pairwise distance across the matched sample after fitting and using logistic regression as the distance function. |
| Study size | 10 | Explain how the study size was arrived at (if applicable) | 99.4% of patients who received MAb were included in the post-PS analysis. This cohort was chosen to allow at least 30 days follow up during a period (6/1/2021-1/31/2022) dominated by SARS-CoV-2 Delta and Omicron subvariants. |

| | | | |
|---|---|---|---|
| Quantitative variables | 11 | Explain how quantitative variables were handled in the analyses. If applicable, describe which groupings were chosen and why | Both continuous variables and parameterization of categorical variables are explained Method section. |
| Statistical methods | 12 | (*a*) Describe all statistical methods, including those used to control for confounding | The Methods section articulate our optimal propensity matching method, exact McNemar's test comparing the proportions in the pair dataset and 95% confidence intervals. |
| | | (*b*) Describe any methods used to examine subgroups and interactions | We also fitted a multivariable Cox proportional hazard regression model predicting the composite outcome in the PS matched subgroups. The Kaplan-Meier estimator was used to plot curves for the composite outcome between the post-PS matched groups. |
| | | (*c*) Explain how missing data were addressed | The vaccination status was missing for 13.9% of the patients in the post-propensity matched cohort (Table 1). Missing vaccination status was analyzed as a separate category (fully vaccinated, not fully vaccinated, unknown (missing). |
| | | (*d*) *Cohort study*—If applicable, explain how loss to follow-up was addressed | We assumed that all patients in the study cohort were followed in the Banner Healthcare system and follow-up was complete. We stated the possibility of primary outcome out-of-Banner system hospitalization as a limitation in the Discussion section. |

| | | | |
|---|---|---|---|
| | | *Case-control study*—If applicable, explain how matching of cases and controls was addressed | *N/A* |
| | | *Cross-sectional study*—If applicable, describe analytical methods taking account of sampling strategy | |
| | | (*e*) Describe any sensitivity analyses | *N/A* |
| **Results** | | | |
| Participants | 13* | (a) Report numbers of individuals at each stage of study—eg numbers potentially eligible, examined for eligibility, confirmed eligible, included in the study, completing follow-up, and analyzed | See Flow Chart (Figure 1). |
| | | (c) **Use of a flow diagram** | See Flow Chart (Figure 1) |
| Descriptive data | 14* | (a) Give characteristics of study participants (eg demographic, clinical, social) and information on exposures and potential confounders | Tables 1 include summary statistics of both exposures and confounders. |

| | | | |
|---|---|---|---|
| | | (b) Indicate number of participants with missing data for each variable of interest | Included in Table 1 |
| | | (c) *Cohort study*—Summarise follow-up time (eg, average and total amount) | Included in Table 1. Each patient had at least 30 days follow-up post-index date. |
| Outcome data | 15* | *Cohort study*—Report numbers of outcome events or summary measures over time | Included in Table 2 and Table3. |
| | | *Case-control study*—Report numbers in each exposure category, or summary measures of exposure | *N/A* |
| | | *Cross-sectional study*—Report numbers of outcome events or summary measures | *N/A* |
| Main results | 16 | (*a*) Give unadjusted estimates and, if applicable, confounder-adjusted estimates and their precision (eg, 95% confidence interval). Make clear which confounders were adjusted for and why they were included | Comparative proportions along with confidence intervals in the post-propensity score matched cohort and their re-matched subgroups in Table 2-3. |
| Other analyses | 17 | Report other analyses done—eg analyses of subgroups and interactions, and sensitivity analyses | Subgroup analysis for vaccinated status was described and shown in Table 3. Sensitivity analysis was not performed and interaction effects were not tested. |
| **Discussion** | | | |
| Key results | 18 | Summarise key results with reference to study objectives | Description of results aligns with stated objective of estimating independent effects of the Cas-Imd and Sotrovimab MAb use. |

| | | | |
|---|---|---|---|
| Limitations | 19 | Discuss limitations of the study, taking into account sources of potential bias or imprecision. Discuss both direction and magnitude of any potential bias | Limitations, including retrospective study design, unmeasured confounding, prior COVID -19 exposures, lack of viral genotyping, and others are discussed extensively in the Discussion section. |
| Interpretation | 20 | Give a cautious overall interpretation of results considering objectives, limitations, multiplicity of analyses, results from similar studies, and other relevant evidence | Caution is incorporated into the discussion and detailed in the limitations section. |
| Generalisability | 21 | Discuss the generalisability (external validity) of the study results | Results are clearly framed as being generalizable only to Southwestern U.S. Further real-world research from large healthcare organizations in different regions of the U.S. would be needed to assess generalizability. |

*Give information separately for cases and controls in case-control studies and, if applicable, for exposed and unexposed groups in cohort and cross-sectional studies.

**Note:** An Explanation and Elaboration article discusses each checklist item and gives methodological background and published examples of transparent reporting. The STROBE checklist is best used in conjunction with this article (freely available on the Web sites of PLoS Medicine at http://www.plosmedicine.org/, Annals of Internal Medicine at http://www.annals.org/, and Epidemiology at http://www.epidem.com/). Information on the STROBE Initiative is available at www.strobe-statement.org.
