## Supplemental Document for "EFFECTIVENESS OF CASIRIVIMAB-IMDEVIMAB AND SOTROVIMAB MONOCLONAL ANTIBODY TREATMENT AMONG HIGH-RISK PATIENTS WITH SARS-CoV-2 INFECTION: A REAL-WORLD EXPERIENCE"

Supplemental Document A:

Website for the Banner Health COVID-19 Treatment:

https://www.bannerhealth.com/staying-well/health-and-wellness/wellness/covid/treatment

Supplemental Table:

Supplemental Table S1. The highest level of oxygen requirement among the post-propensity matched hospitalized patients within 30 days of the index date.

|  | mAb | Control | P-value | Cas-Imd | Control | P-value | Sotrovimab | Control | P-value |
| --- | --- | --- | --- | --- | --- | --- | --- | --- | --- |
|  | N=439 | N=849 |  | N=401 | N=800 |  | N=38 | N=49 |  |
| **O_2_ Therapy** |  |  | <0.01* |  |  | <0.01* |  |  | 0.67* |
| Mechanical ventilation and non-invasive ventilation (CPAP-BiPAP/ high flow O_2_) | 28 (6.4) | 116 (13.7) |  | 26 (6.5) | 114 (14.2) |  | 2 (5.3) | 2 (4.1) |  |
| Nasal Cannula | 118 (26.9) | 294 (34.6) |  | 113 (28.2) | 284 (35.5) |  | 5 (13.2) | 10 (20.4) |  |
| No Oxygen Therapy | 293 (66.7) | 439 (51.7) |  | 262 (65.3) | 402 (50.2) |  | 31 (81.6) | 37 (75.5) |  |
| Fully Vaccinated** | 64 (14.6) | 127 (15.0) | 0.47 | 51 (12.7) | 112 (14.0) | 0.33 | 13 (34.2) | 15 (30.6) | 0.89 |
| Admission to ICU** | 57 (13.0) | 167 (19.7) | <0.01 | 51 (12.7) | 158 (19.8) | <0.01 | 6 (15.8) | 9 (18.4) | 0.98 |

Data are presented as mean [SD] for continuous measures, and n (%) for categorical measures.

Abbreviations: Cas-Imd: casirivimab-imdevimab; CPAP: continuous positive airway pressure; BiPAP: bilevel positive airway pressure; ICU: intensive care unit.

*p-values account for pairing and are based on a Wald test.

**Vaccination status and admission to ICU p-values do not account for pairing and showing the results of chi-squared testing.

Supplemental Figures:

Supplemental Figure S1. Duration between COVID-19 test positivity and monoclonal antibody infusion among casirivimab-imdevimab and sotrovimab cohorts (days).

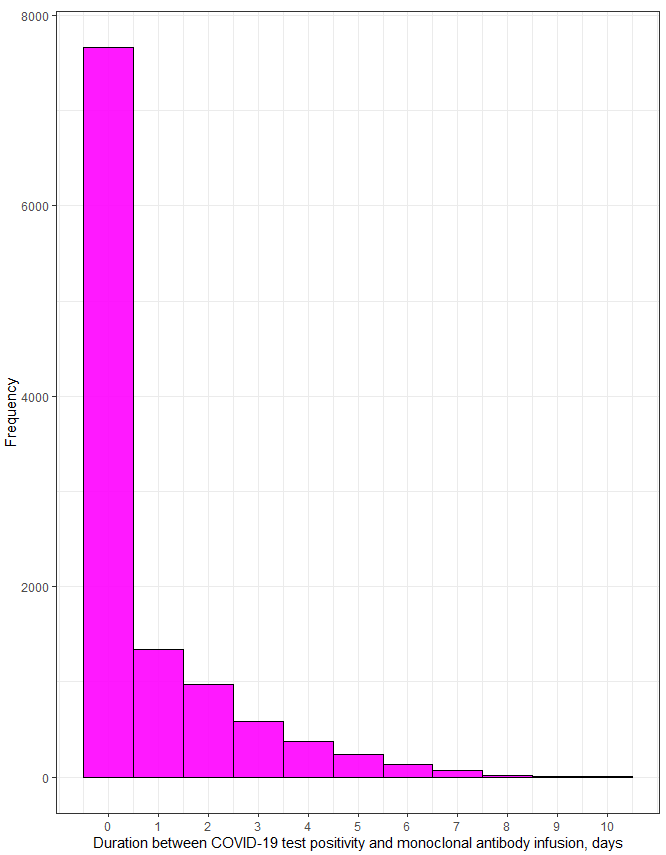

Supplemental Figure S2. Duration between COVID-19 test positivity and monoclonal antibody infusion among casirivimab-imdevimab monoclonal antibody cohort (days).

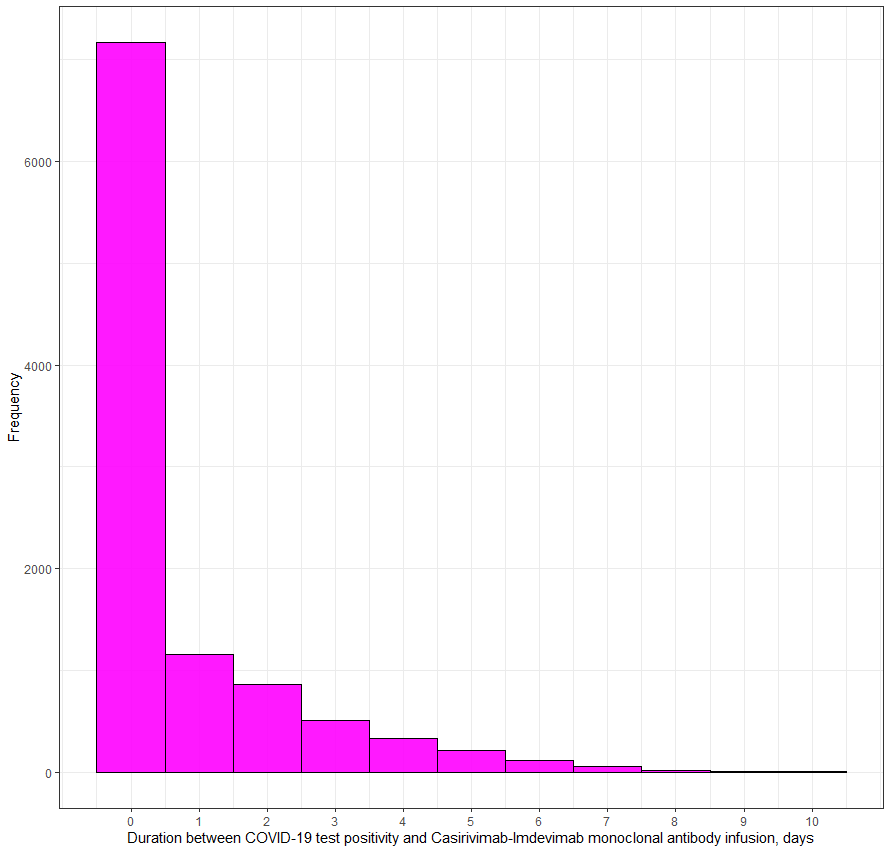

Supplemental Figure 3. Duration between COVID-19 test positivity and monoclonal antibody infusion among sotrovimab monoclonal antibody cohort (days).

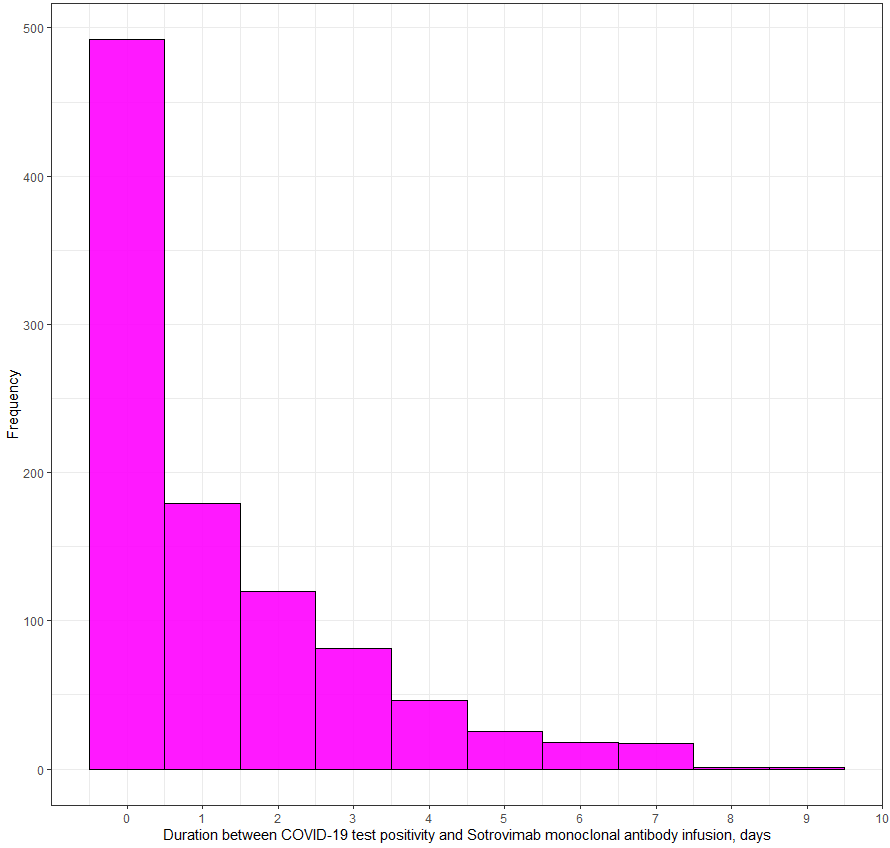

Supplemental Figure S4. Kaplan-Meier survival curves for the effect of monoclonal antibody administration on the composite outcome compared to the post-propensity matched untreated control cohort.

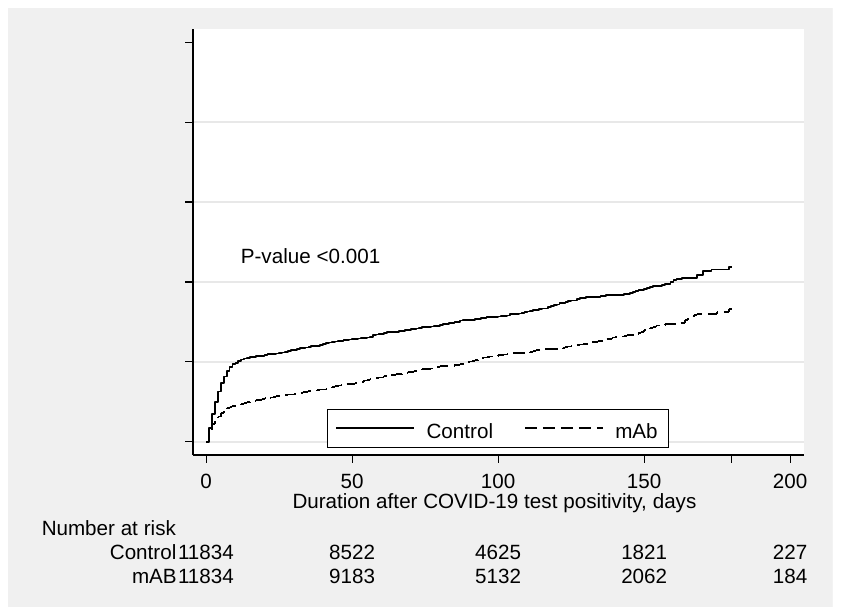

Supplemental Figure S5. Kaplan-Meier survival curves for the effect of Casirivimab-Imdevimab monoclonal antibody administration on the composite outcome compared to the post-propensity matched untreated control cohort.

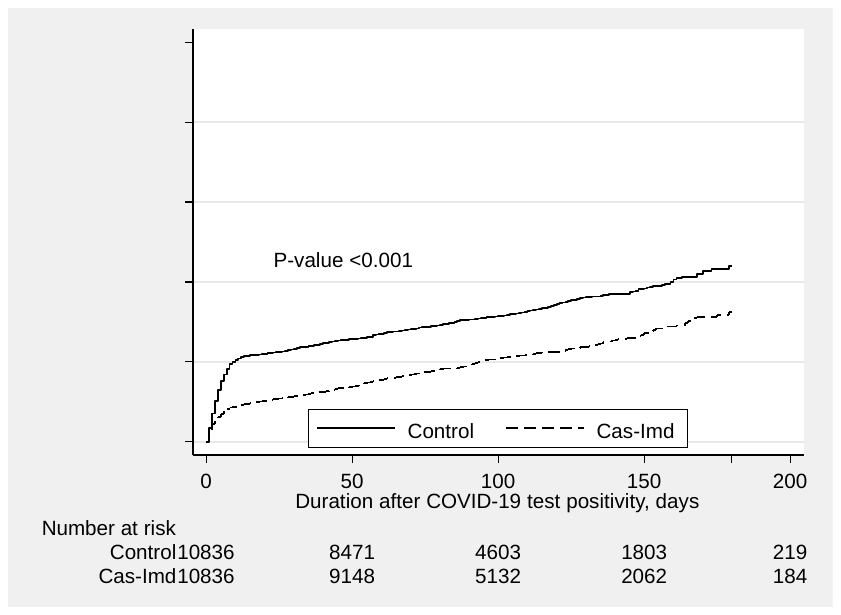

Supplemental Figure S6. Kaplan-Meier survival curves for the effect of sotrovimab monoclonal antibody administration on the composite outcome compared to the post-propensity matched untreated control cohort.

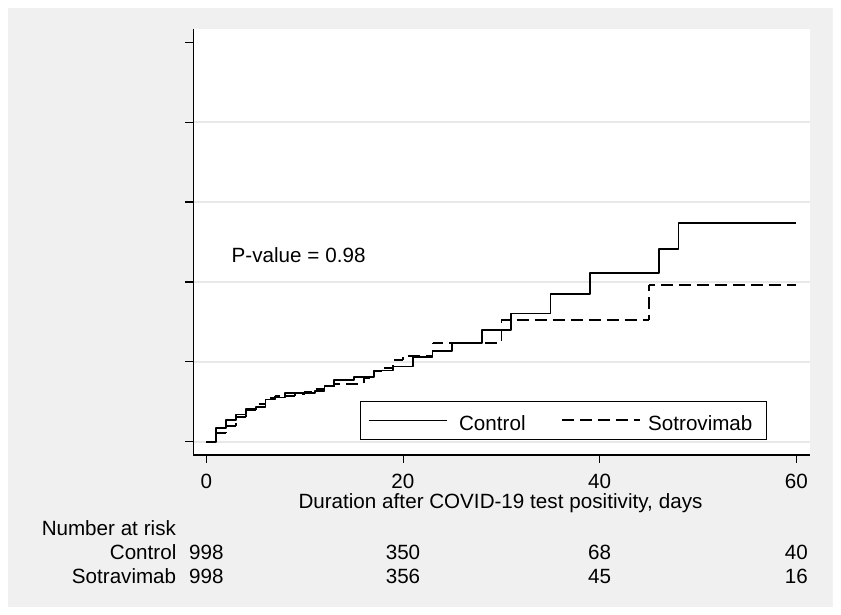
